# Neuropathy Assessment and Treatment Patterns in Patients with Hereditary Transthyretin Amyloidosis

**DOI:** 10.64898/2026.04.15.26350949

**Authors:** Nicholas Streicher, Hermela Wubet

**Author notes:** Corresponding author: Nicholas Streicher, MD, Department of Neurology, Georgetown University, Washington, DC.

## Abstract

**Background and Objectives:** Hereditary transthyretin amyloidosis (hATTR) manifests as cardiomyopathy and/or polyneuropathy. Patients may be thought to have neuropathy for a variety of reasons, but the clinical evidence supporting that impression varies. We used ICD-10 coding to broadly capture patients thought to have neuropathy in a V142I-predominant cohort and examined what clinical documentation actually supported the diagnosis and whether documentation level influences treatment selection.

**Methods:** Retrospective chart review of 54 patients identified by co-occurring ICD-10 codes for hereditary transthyretin amyloidosis and polyneuropathy at a major academic medical center, with pathogenic TTR variant confirmation. Neuropathy codes were then classified by the level of clinical evidence supporting the diagnosis. Treatment with TTR stabilizers and gene silencers was assessed.

**Results:** Of 54 patients (88.9% African American, 85.2% V142I), 51 (94.4%) had confirmed cardiac involvement with 40/42 eligible (95.2%) receiving stabilizers. Sixteen patients (29.6%) received gene silencers, with 13 receiving both concurrently. Clinical evidence supporting the neuropathy code was identified in 30 patients (55.6%): ancillary testing confirmation in 17 (31.5%), provider documentation without ancillary testing in 13 (24.1%), and symptoms only in 10 (18.5%). The remaining 14 (25.9%) had no clear clinical basis for the code. Gene silencer use was highest among those with ancillary testing (47.1%) versus symptoms only (10.0%).

**Discussion:** Among hATTR patients coded for neuropathy, only 31.5% had ancillary testing confirmation. Gene silencer use tracked with documentation quality rather than the presence of a code alone. These findings highlight the gap between administrative coding and clinical documentation and support standardized neurological assessment in hATTR.

## INTRODUCTION

Hereditary transthyretin amyloidosis (hATTR) is an autosomal dominant systemic disease caused by mutations in the transthyretin (TTR) gene that lead to misfolding and tissue deposition of amyloid fibrils.^1,13^ Clinical manifestations primarily affect the heart and peripheral nervous system, with phenotypic expression varying by specific genetic variant, geographic ancestry, and individual factors.^2,13^

The V142I variant is the most common pathogenic TTR mutation worldwide, carried by approximately 3–4% of African Americans, representing an estimated 1.2 to 1.5 million individuals in the United States.^2,3^ This variant classically produces a cardiac-predominant phenotype with heart failure symptoms typically manifesting after age 60, leading to its historical characterization as a cardiomyopathy.^2^ However, emerging evidence suggests neurological involvement may be more prevalent than previously recognized, with large database studies demonstrating significantly elevated polyneuropathy diagnoses among V142I carriers compared to non-carriers.^4,5^

The therapeutic landscape for hATTR has expanded substantially. TTR stabilizers (tafamidis, acoramidis, diflunisal) prevent tetramer dissociation, with tafamidis demonstrating mortality benefit in ATTR cardiomyopathy^6^ and evidence supporting neurological benefit in patients with mixed phenotypes.^7,8^ Gene silencer therapies (patisiran, vutrisiran, eplontersen) reduce hepatic TTR synthesis through RNA interference or antisense mechanisms, with established efficacy for polyneuropathy^9,10,11^ and emerging data supporting cardiac benefits.^12^ Understanding treatment utilization patterns, particularly for gene silencers that may address both cardiac and neurological manifestations, has important implications as therapeutic indications evolve.

We conducted a retrospective analysis of patients with co-occurring ICD-10 codes for hereditary transthyretin amyloidosis and polyneuropathy at a major academic medical center. Our aim was to examine what clinical evidence supports neuropathy coding in this population, whether documentation quality varies across patients, and whether the level of neuropathy documentation influences treatment selection, particularly gene silencer utilization, in this predominantly V142I cohort.

## METHODS

### Study Design and Population

We performed a retrospective cohort study of patients diagnosed with hATTR at a major academic medical center. Patients were identified through institutional electronic medical record query of encounters over approximately eight years (2017–2025), using ICD-10 coding for amyloidosis and neuropathy diagnoses, with confirmation of a pathogenic TTR variant through genetic testing documentation. All cases were individually reviewed to verify clinical status. No patients in our cohort were prescribed gene silencers under the ATTR cardiomyopathy indication; all gene silencer use reflected the polyneuropathy indication. We excluded patients with unknown genotype, those receiving hospice or comfort care only, and pre-symptomatic carriers identified through family screening who had not developed clinical manifestations. The Institutional Review Board of MedStar Health Research Institute gave ethical approval for this work. Written informed patient consent was waived due to the retrospective nature of the study.

### Data Collection

Electronic medical records were reviewed for demographic information, TTR variant, cardiac involvement documentation (echocardiography, cardiac MRI, technetium pyrophosphate scintigraphy), neurological assessments (neurology consultations, electromyography/nerve conduction studies), and treatments received.

### Neuropathy Classification

Patients were classified into four mutually exclusive categories based on highest level of neuropathy documentation:

> *Neuropathy with ancillary testing*: abnormal EMG/nerve conduction study consistent with polyneuropathy.
>
> *Neuropathy without ancillary testing*: clinical documentation of neuropathy by a provider without confirmatory ancillary testing.
>
> *Symptoms only*: patient-reported neuropathic symptoms (numbness, tingling, pain) documented in medical records without clinical examination findings or formal neurological evaluation.
>
> *Unclear*: insufficient documentation to assess neurological status.

“Confirmed or suspected neuropathy” was defined as the combined group of patients with neuropathy with ancillary testing, neuropathy without ancillary testing, or symptoms only, representing all patients with any clinical evidence suggesting neurological involvement.

### Treatment Assessment

Current or prior treatment was assessed for TTR stabilizers (tafamidis, acoramidis, diflunisal) and gene silencers (patisiran, vutrisiran, eplontersen). Stabilizer eligibility for cardiac patients was defined as confirmed ATTR cardiomyopathy without documented contraindication or patient refusal.

### Confounding Conditions

We documented conditions that may cause or contribute to neuropathy independent of amyloidosis: diabetes mellitus, chronic kidney disease (eGFR <60), alcohol use disorder, and vitamin B12 deficiency. These confounders were assessed to contextualize neuropathy findings and treatment decisions.

## RESULTS

### Patient Characteristics

Fifty-four patients met inclusion criteria (Table 1). The cohort was predominantly African American (48/54, 88.9%) with V142I as the most common variant (46/54, 85.2%). Other variants included V50M (3 patients), A140S (2 patients), T60A, V132I, and P84L (1 patient each). Median age at diagnosis was 72 years (range 51–89).

**Table 1.** Patient Demographics and Clinical Characteristics (N=54)

| Characteristic | n (%) |
| --- | --- |
| **Race** |  |
| African American | 48 (88.9) |
| White | 5 (9.3) |
| Unknown | 1 (1.9) |
| **TTR Variant** |  |
| V142I | 46 (85.2) |
| V50M | 3 (5.6) |
| A140S | 2 (3.7) |
| Other (T60A, V132I, P84L) | 3 (5.6) |
| **Cardiac Status** |  |
| Confirmed cardiac involvement | 51 (94.4) |
| No confirmed cardiac involvement | 3 (5.6) |
| **Treatment** |  |
| Stabilizer | 40 (74.1) |
| Gene silencer | 16 (29.6) |
| Both stabilizer + gene silencer | 13 (24.1) |

Values may not add to 100% because of rounding.

Confirmed cardiac involvement was present in 51 patients (94.4%), consistent with the expected cardiac-predominant phenotype of V142I.^2^ Among patients with confirmed cardiomyopathy who were eligible for stabilizer therapy, 40 of 42 (95.2%) were receiving treatment. Thirteen patients (24.1%) were receiving both a stabilizer and a gene silencer concurrently, with gene silencers added to stabilizer therapy rather than replacing it. Nine were ineligible due to contraindication or refusal, and two eligible patients were not receiving stabilizers for reasons not clearly ascertainable from records.

### Neuropathy Classification

Neuropathy classification and treatment patterns are shown in Table 2. Neuropathy confirmed by ancillary testing (abnormal EMG in this cohort) was documented in 17 patients (31.5%), neuropathy without ancillary testing in 13 (24.1%), and symptoms only in 10 (18.5%). Fourteen patients (25.9%) had unclear neurological status. Overall, 30 patients (55.6%) had confirmed or suspected neuropathy based on available documentation.

**Table 2.**
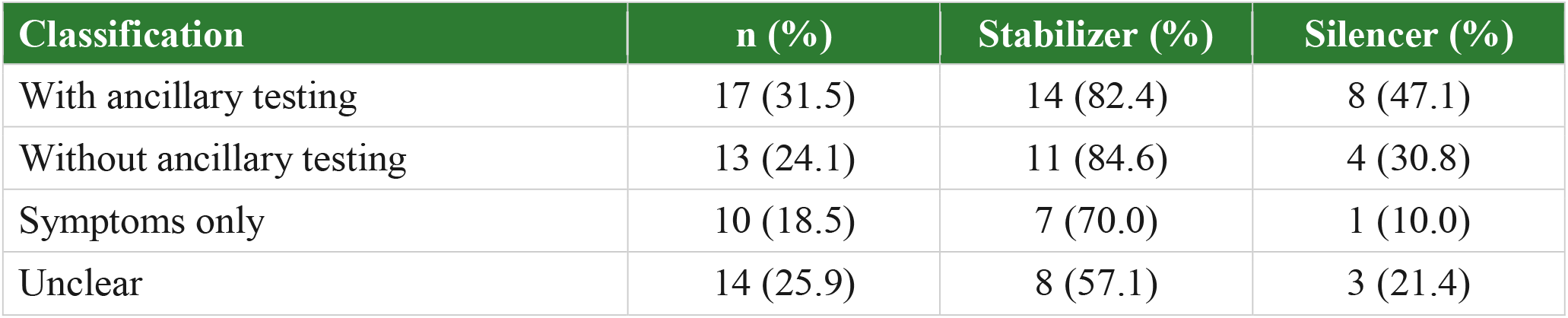
Neuropathy Classification and Treatment Patterns (N=54)

### Confounding Conditions

Table 2a shows treatment patterns stratified by presence of confounding conditions. Among the 16 patients without confounders, 13 (81.2%) received stabilizers and 5 (31.2%) received gene silencers. Among 38 patients with one or more confounders, 27 (71.1%) received stabilizers and 11 (28.9%) received gene silencers.

**Table 2a.** Treatment by Presence of Confounding Conditions (N=54)

| Confounder Status | n | Stabilizer (%) | Silencer (%) |
| --- | --- | --- | --- |
| No confounders | 16 | 13 (81.2) | 5 (31.2) |
| With confounders | 38 | 27 (71.1) | 11 (28.9) |

Confounders include: diabetes mellitus (DM), chronic kidney disease (CKD; eGFR <60), alcohol use disorder (EtOH), and vitamin B12 deficiency.

### Analysis by Cardiac Status

Table 3 presents neurological assessment and treatment stratified by cardiac status. Among 51 patients with cardiac involvement, 29 (56.9%) had confirmed or suspected neuropathy. Neurology consultation rates were higher among those with confirmed or suspected neuropathy (28/29, 96.6%) compared to unclear status (15/22, 68.2%). EMG completion was 75.9% among those with confirmed or suspected neuropathy versus 40.9% in the unclear group.

**Table 3.** Neurological Assessment and Treatment by Cardiac Status.

| Cardiac Involvement | Neuropathy Classification | n | Neurology Consultation (%) | EMG (%) | Silencer (%) | Stabilizer (%) |
| --- | --- | --- | --- | --- | --- | --- |
| Yes (n=51) | Confirmed/Suspected | 29 | 28 (96.6) | 22 (75.9) | 11 (37.9) | 25 (86.2) |
|  | Unclear | 22 | 15 (68.2) | 9 (40.9) | 2 (9.1) | 15 (68.2) |
| No (n=3) | Confirmed/Suspected | 1 | 1 (100) | 1 (100) | 1 (100) | 0 (0) |
|  | Unclear | 2 | 2 (100) | 2 (100) | 2 (100) | 0 (0) |

Three patients (one V142I, one V50M, one P84L) did not have confirmed cardiac involvement. All three received gene silencers rather than stabilizers, consistent with a non-cardiac phenotype, although documentation surrounding neuropathy varied.

## DISCUSSION

### V142I Phenotype and Neurological Involvement

This single-center analysis of 54 hATTR patients, predominantly V142I carriers of African ancestry, confirms the expected cardiac-predominant phenotype while revealing substantial rates of confirmed or suspected neuropathy. The finding that 55.6% (30/54) had documented neuropathic symptoms or signs aligns with recent population-based studies demonstrating elevated polyneuropathy prevalence among V142I carriers,^4^ challenging the traditional characterization of this variant as exclusively cardiac.^5^ Notably, reported neuropathy prevalence among V142I carriers varies widely across studies, from approximately 2–9% using administrative ICD-10 coding^4^ to over 90% at specialized centers employing nerve conduction studies and skin biopsy^5^, highlighting that diagnostic methodology may be a primary driver of apparent variation in disease prevalence.

Beyond methodology, confounding conditions complicate interpretation. The majority of patients had at least one condition independently associated with neuropathy, including diabetes mellitus, chronic kidney disease, alcohol use disorder, and vitamin B12 deficiency. Neuropathy in a given patient may not be attributable to amyloid deposition alone. Both the method of assessment and the burden of confounders must be considered when interpreting neuropathy prevalence in hATTR.

Our observation that three patients lacked confirmed cardiac involvement despite carrying pathogenic TTR variants (one V142I, one V50M, one P84L) is noteworthy. These patients may represent early detection through cascade screening or genuinely neurologic-predominant phenotypes. As genetic testing expands, identifying gene-positive individuals before cardiac manifestations develop will become more common.

### Treatment Patterns

Stabilizer utilization among eligible cardiac patients was high (95.2%), reflecting established practice following pivotal trial data demonstrating mortality benefit.^6^ Gene silencer use showed a clear gradient by neuropathy documentation: highest in patients with ancillary testing confirmation (47.1%), intermediate with neuropathy documented without ancillary testing (30.8%), and lowest with symptoms only (10.0%). Notably, 13 of 16 patients receiving gene silencers were also on concurrent stabilizer therapy, reflecting an additive prescribing pattern in which gene silencers were layered onto existing stabilizer treatment rather than substituted for it.

Three patients without confirmed cardiac disease received gene silencers, consistent with a neurologic-predominant phenotype. No patients in our cohort were prescribed gene silencers under the recently approved ATTR cardiomyopathy indication (FDA March 2025, based on HELIOS-B trial data^12^); all gene silencer access was predicated on neurological documentation.

With vutrisiran now approved for cardiomyopathy, cardiologists can prescribe gene silencers without neurological documentation. Neuropathy may go uncharacterized as its documentation is no longer required for treatment access.

Our finding that over half of patients in a cardiac-predominant cohort had documented neuropathy suggests that systematic neurological assessment adds clinical value beyond treatment access, including establishing baseline neuropathy severity, guiding monitoring, and identifying patients who may benefit from multidisciplinary management.

### Limitations

This retrospective single-center analysis relies on available documentation. No patients in our cohort were prescribed gene silencers under the ATTR cardiomyopathy indication (FDA approved March 2025); all gene silencer use reflects the polyneuropathy indication, and current prescribing patterns may differ as adoption of the expanded indication increases.

Our V142I-predominant cohort limits generalizability to other variants. The small number of patients without cardiac involvement (n=3) limits statistical inference for that subgroup. These patients may represent early detection through cascade screening or genuinely neurologic-predominant phenotypes, a pattern likely to become more common as genetic testing expands.

As a retrospective chart review relying on administrative coding for cohort identification, ICD-10 codes may overcapture or undercapture patients. All cases were individually reviewed, leading to the exclusion of four identified carriers. Ancillary neuropathy testing was limited to EMG/nerve conduction studies; skin biopsy and autonomic testing were not performed. We could not assess clinical reasoning behind individual treatment decisions, including patient preferences and insurance barriers.^5^

### Future Directions

The wide variation in reported neuropathy prevalence among V142I carriers, from approximately 2–9% in population-based studies to over 90% at specialized centers, likely reflects differences in diagnostic methodology. Emerging tools including serum neurofilament light chain (NfL), nerve ultrasound, and structured patient-reported outcome measures may help standardize detection and provide perspectives on the diagnostic journey not captured in chart review.

Our own data illustrate this: the apparent neuropathy rate in our cohort ranges from 31.5% when restricted to objective EMG confirmation to 55.6% when clinical documentation of symptoms is included. Standardizing neuropathy assessment criteria in hATTR, including which diagnostic modalities to employ and how to account for competing etiologies in this patient population, represents an important area for future investigation.

## CONCLUSIONS

In this predominantly V142I cohort, 94.4% had confirmed cardiac involvement, consistent with the known cardiac-predominant phenotype. Stabilizer use was high (95.2%) among eligible cardiac patients. Over half (55.6%) had confirmed or suspected neuropathy based on clinical documentation, though EMG completion remained limited. Gene silencer utilization was highest among patients with ancillary testing confirmation of neuropathy and those without cardiac involvement. These findings support systematic neurological assessment in hATTR even when cardiac disease predominates. Both the method of neuropathy diagnosis and the burden of confounding conditions influence apparent prevalence and complicate attribution. While this study characterizes the gap between neuropathy coding and clinical documentation, important questions remain regarding optimal diagnostic modalities, the role of confounders, and how evolving treatment indications will reshape the relationship between neurological assessment and treatment access in hATTR.

## Data Availability

Data is protected by HIPPA

